# Effects of an educational program on midwives’ intentions to prevent premature birth: A randomized controlled trial

**DOI:** 10.1101/2025.10.08.25337616

**Authors:** Masaki Kidera, Shoko Takeuchi, Eriko Shinohara, Kazumi Kubota, Sachiyo Nakamura

**Affiliations:** Yokohama City University, Graduate School of Medicine, Department of Nursing, Yokohama, Japan; Shimonoseki City University, Faculty of Data Science Department of Data Science, Shimonoseki, Japan

**Keywords:** Premature Birth, Midwifery, Randomized Controlled Trial, Evidence-Based Practice

## Abstract

**Background:** Premature birth is a leading cause of neonatal mortality and long-term disability worldwide. Midwives in Japan have few opportunities to receive systematic education on premature birth prevention. We developed an online interactive midwifery education program to enhance midwives’ intention, knowledge, and attitudes toward evidence-based practice (EBP) for premature birth prevention.

**Methods:** We conducted a randomized controlled trial with a 1:1 allocation ratio. A total of 158 midwives were randomly assigned to either the intervention group (IG; n = 79) or the control group (CG; n = 79). The IG received a 120-minute online interactive education program, whereas the CG received an informational pamphlet. The primary outcome was the intention to practice midwifery care for premature birth prevention two weeks after intervention, assessed using the Continuing Professional Development–Reaction Questionnaire. Secondary outcomes included knowledge and attitudes toward EBP, measured using the Evidence-Based Practice Attitude Scale (EBPAS). Due to the nature of the intervention, blinding of participants and researchers was not feasible; allocation concealment was maintained.

**Results:** All randomized participants were included in the analysis. The median intention score was significantly higher in the IG than in the CG immediately after the program (6.5 [IQR: 5.5– 7.0] vs. 5.5 [IQR: 5.0–6.5]; p = .005, effect size = 0.23 [small]) and two weeks after the intervention (6.0 [IQR: 5.5–7.0] vs. 5.5 [IQR: 5.0–6.0]; p = .001, effect size = 0.27 [small]). Knowledge scores were also significantly higher in the IG both immediately after (9.0 [IQR: 9.0–10.0] vs. 6.0 [IQR: 6.0–7.0]; p < .001, effect size = 0.81[large]) and two weeks after the program (9.0 [IQR: 8.0–10.0] vs. 6.0 [IQR: 4.3–7.0]; p < .001, effect size = 0.72 [large]). No significant differences between groups were observed in EBPAS scores at any time point.

**Conclusions:** The online interactive education program effectively improved midwives’ intention and knowledge regarding premature birth prevention.

**Trial Registration:** University Hospital Medical Information Network Clinical Trials Registry (UMIN-CTR): UMIN000058251.

**Funding:** This study was supported by the Yamaji Fumiko Specialized Nursing Education Research Grant Fund.

## Introduction

Premature birth—defined as delivery before 37 completed weeks of gestation—is a leading cause of perinatal morbidity, mortality, and long-term disability [1, 2]. The complications of premature birth can have devastating and lifelong consequences for newborns and children [2] and represent the primary cause of death among children under five years of age [3]. In Japan, the premature birth rate is approximately 5.7% [4], which is lower than the global range of 4–16% [3]. However, the consequences of premature birth extend beyond infants, affecting families, caregivers, healthcare systems, and society at large [5]. Reducing premature births is thus an urgent public health priority.

The causes of premature birth are multifactorial, encompassing maternal and fetal conditions such as gestational diabetes and infection [6], as well as behavioral and environmental risk factors including smoking [7], poor nutrition [8], folic acid deficiency [9], stress [10], and periodontal disease [11]. Because lifestyle, environmental, and psychosocial factors influence risk, comprehensive support during pregnancy is essential. The World Health Organization [12] recommends promoting healthy pregnancies through counseling on nutrition, smoking cessation, and avoidance of harmful substances.

In Japan, midwives have traditionally provided guidance to support healthy pregnancies. Evidence-based maternity care guidelines were first published in 2012 and revised in 2024 [13]. However, preventive efforts remain insufficient. For example, inadequate gestational weight gain is associated with an increased risk of premature birth [14], yet 66% of women reported receiving no guidance on weight gain from healthcare professionals [15]. Moreover, Intimate Partner Violence (IPV) is linked to premature birth [16]. Although national guidelines recommend routine IPV screening during pregnancy [17], only 6.2% of facilities conduct such screening appropriately [18].

Implementing Evidence-Based Practice (EBP) can improve maternal, fetal, and newborn outcomes [19]. However, barriers to EBP among nurses and midwives include limited knowledge, skills, motivation, and resources [20]. Therefore, the strategies to promote EBP must address both professional competence and motivation for behavioral change. According to Theory of Planned Behavior (TPB), attitudes, subjective norms, and perceived behavioral control shape intentions, which in turn predict behavior [21].

Educational interventions based on the TPB have been shown to improve nurses’ care for IPV survivors by enhancing attitudes, norms, and perceived control [22]. Furthermore, a systematic review found the TPB to be the strongest predictor of healthcare professionals’ clinical behaviors [23]. Given its effectiveness in promoting behavioral change, we developed an online interactive education program grounded in the TPB to promote premature birth prevention among midwives. The primary aim of this study was to evaluate the effect of this program on midwives’ intentions to adopt preventive care practices.

### Research hypotheses

The hypotheses of this study are as follows:

H1: The intention to practice midwifery care to prevent premature birth immediately after and two weeks after the intervention will be significantly higher in the intervention group (IG) than in the control group (CG).

H2: The knowledge scores regarding midwifery care to prevent premature birth immediately after and two weeks after the intervention will be significantly higher in the IG than in the CG.

H3: Evidence-Based Practice Attitude Scale (EBPAS) scores immediately after and two weeks after the intervention will be significantly higher in the IG than in the CG.

## Materials and methods

### Trial design

This study employed a randomized controlled trial (RCT) design to compare the educational effects of the program between the intervention group (IG) and the control group (CG). The study was conducted during the summer of 2024 and adhered to the Consolidated Standards of Reporting Trials (CONSORT) guidelines for reporting randomized trials [24]. Data were collected between September 2024 and March 2025.

### Participants and recruitment

Participants were recruited through a midwifery career website. A study announcement was posted on the website, and midwives who were interested accessed an online study information sheet. Those who agreed to participate submitted an application form via the website. After confirming their eligibility, the researchers mailed written informed consent forms to the applicants. Recruitment was conducted from October 8, 2024, to February 28, 2025.

Before the start of the educational program, an online meeting was held to provide participants with a detailed explanation of the study procedures, potential risks and benefits, and ethical considerations. Midwives who wished to participate signed the consent form and returned it by mail. They were informed that participation was voluntary, that they could withdraw from the study at any time without penalty, and that a withdrawal form was included along with the consent form.

The inclusion criteria were as follows: (a) being a licensed midwife and (b) having the ability to participate in educational programs via the Internet.

### Sample size

This study used the statistical software G-Power version 3.1.9.7. for calculations. For the calculation of the sample size, this study referred to previous studies [22] related to the TPB to set the effect size at 0.47. The sample size for this study was determined based on a power analysis conducted with an α error of 0.05 and a statistical power of 0.8. Based on these calculations, 72 midwives were required for the IG and 72 for the CG. Assuming a 10% dropout rate, 158 midwives were recruited for the intervention and control groups.

### Randomization, allocation concealment, and blinding

After written informed consent was obtained, participants were randomized 1:1 to the intervention or control group using a computer-generated sequence prepared and safeguarded by an independent statistician who had no role in participant enrollment, intervention delivery, or outcome assessment. Because blinding of participants and study personnel was not feasible—the data collectors also delivered the educational sessions— masking was not applied. In addition, outcome assessors were not blinded because the same researchers who delivered the intervention also collected and analyzed the data. To mitigate potential bias inherent to the unblinded design, allocation concealment was implemented: enrollment staff had no access to the randomization list, and group assignments were disclosed only after completion of the consent procedures. The full study protocol is available as Supplementary File S1 Protocol.

### Development of the educational program

#### Program objectives and structure

This program was designed to increase midwives’ intentions to engage in midwifery care practices for premature birth prevention, as intention is a prerequisite for behavior. The program consisted of lectures, practical training, and group discussions, with a total duration of 120 minutes (Table 1). Participants were given a pre-task to reflect on the midwifery care they had provided for premature birth prevention and a post-task to consider the care they would like to implement in the future.

**Table. 1.** Education program structure.

| Learning contents | Learning objects | TPB contents | Time |
| --- | --- | --- | --- |
| 1.Pre-assignment | • Reflect on the antenatal care you are currently practicing. | Attitude |  |
| 2. Presentation of learning objects | • Preventing premature birth by improving the mind, body, and environment of pregnant women |  | 5minutes |
| 3.Group discussion① | • Introducing myself and sharing the pregnancy care I provide. | Attitude | 20minutes |
| 4. Lecture① | • Evidence on midwifery care to prevent preterm birth. | Perceived behavioral control | 25minutes |
|  | • Current situation of preterm birth and the role of midwives. | Social norms |  |
| 5. Lecture② | • The role of midwives in midwifery care was reviewed using the Essential Competencies for Midwifery Practice | Social norms | 25minutes |
| 6. Practice | • Exercises to prevent premature birth with hiesho (sensitivity to cold). | Perceived behavioral control | 20minutes |
| 7. Group discussion② | • About midwifery care to prevent preterm birth that I would like to practice. | Perceived behavioral control | 20minutes |
| 8. Summary | • Lecture summary |  | 5minutes |
| 9. Post-assignment | • Think about the preventive midwifery care you would like to implement within the next two weeks | Attitude |  |

The program was grounded in the Theory of Planned Behavior (TPB) and aimed to enhance intentions by addressing three key determinants: attitudes toward midwifery care for premature birth prevention, subjective norms, and perceived behavioral control (Figure 1). To foster attitude changes, the pre-task encouraged participants to critically reflect on their own midwifery practices. The post-task then prompted them to identify specific care practices they intended to adopt. To strengthen subjective norms, participants reviewed the professional roles and responsibilities of midwives as outlined in the Essential Competencies for Midwifery Practice [25]. To enhance perceived behavioral control, participants were provided with opportunities during the group discussion to consider and exchange ideas about the care they could provide for premature birth prevention.

**Figure 1.**
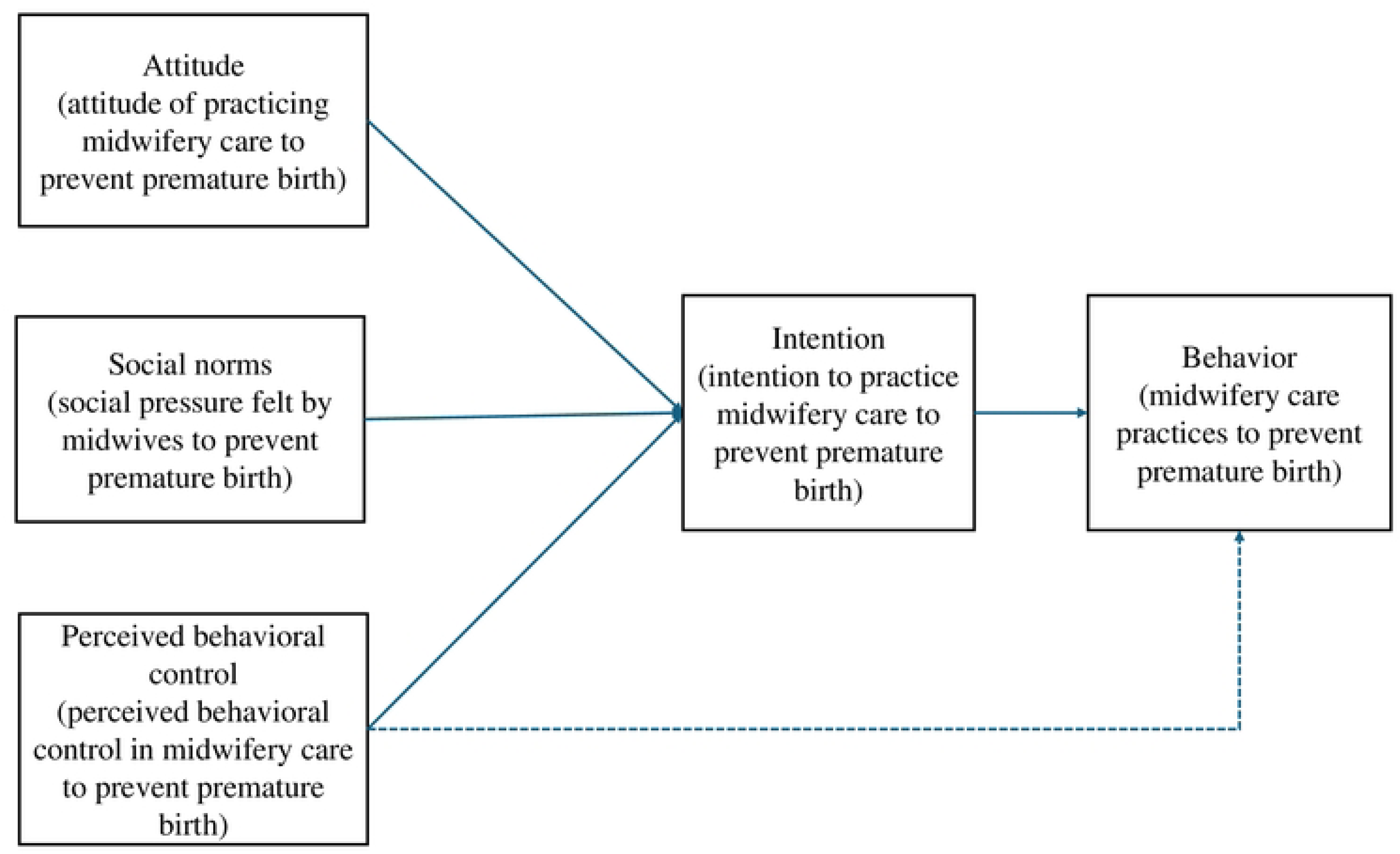
Research framework based on the theory of planned behavior.

In addition, the program addressed hiesho (sensitivity to cold), a condition characterized by cold extremities due to reduced peripheral circulation. Previous studies [26] have reported that pregnant women with hiesho have an approximately 3.4 times higher risk of premature birth. Therefore, light exercises to improve peripheral circulation and alleviate hiesho were incorporated into the practical training component of the program.

The educational program was delivered as live online sessions facilitated by an instructor, enabling participants to actively engage in interactive learning. An online reservation system, developed specifically for the program, offered multiple available dates, from which the participants could select their preferred session. Because participants were free to choose their session dates, the number of attendees varied; however, each session was limited to a maximum of four participants to ensure sufficient interaction. When only one participant attended, the session was conducted as a one-on-one dialogue with the facilitator, allowing the participant to reflect deeply on their own practice. The intervention was implemented by researchers who had substantial clinical experience and were highly skilled in providing health education and counseling for pregnant women. Evidence from a systematic review of nine randomized controlled trials demonstrated that online nursing education is well received and produces more positive effects on self-efficacy in nursing skills compared with traditional face-to-face learning [27]. Furthermore, interactive instructional approaches have been shown to enhance participant satisfaction [28].

#### Development phases

The development of the educational program proceeded in two phases. First, a scoping review was conducted to identify interventions effective in preventing premature birth [29]. Twenty-seven studies met the inclusion criteria. Twelve interventions were identified as effective: nutritional interventions, promotion of smoking cessation, interventions to reduce alcohol and drug intake, exercise interventions, promotion of oral hygiene, management of depression and stress, interventions to improve communication with close contacts, reduction of exposure to passive smoking, addressing violence, and introduction of community resources. These were organized into three domains: “preparing the body,” “preparing the mind,” and “preparing the environment.” The review also suggested that continuous care from early pregnancy delivered by trained caregivers is effective in preventing premature birth. These findings were incorporated into the program.

Second, building on the scoping review, evidence-based interventions for preventing premature birth were extracted from pregnancy care guidelines [30] and the WHO antenatal care recommendations [31] and integrated into the educational content. For example, “preparing the body” includes lectures recommending folic acid supplementation and encouraging protein intake for underweight pregnant women. “Preparing the environment” includes recommendations to routinely screen for IPV during pregnancy. Overall, the program emphasizes that comprehensive interventions addressing the mind, body, and environment are essential for premature birth prevention.

### Intervention

#### Intervention group

The IG participated in a 120-min online interactive midwifery care education program for preventing premature birth.

#### Control group

The CG were informed that they were in the control group and provided with a pamphlet about midwifery continuing care, which was not specific to preterm birth. Participants who wished could attend an educational program after the study ended.

#### Data collection

Data were collected at three time points: (1) baseline (pre-intervention), (2) immediately post-intervention, and (3) two weeks post-intervention. At baseline, participants provided demographic and professional characteristics, including age, years of experience as a midwife, years of antenatal-care experience, possession of advanced midwifery qualifications, current employment setting, and prior training in premature birth prevention.

#### Outcomes

The primary outcome was the intention to practice midwifery care to prevent premature births two weeks after the intervention. Intention was assessed using the Japanese version of the Continuing Professional Development–Reaction Questionnaire, a validated 12-item instrument grounded in the Theory of Planned Behavior that measures changes in health professionals’ clinical behavioral intentions [32]. The questionnaire assesses overall intention and four determinants of intention: (1) social influence (perceived approval or disapproval by significant others), (2) beliefs about capabilities (perceived ability to perform the behavior), (3) moral norms (sense of personal obligation to adopt the behavior), and (4) beliefs about consequences (perceived benefits or harms of the behavior). The instrument was translated into Japanese with permission from the developers and evaluated in a feasibility study, which demonstrated acceptable to excellent internal consistency (Cronbach’s α = 0.73–0.91).

The secondary outcomes were (1) knowledge of midwifery care for preventing premature birth and (2) attitudes toward evidence-based practice. Knowledge was assessed using a 10-item multiple-choice questionnaire (four options per item). Each correct response was awarded one point, yielding a total score ranging from 0 to 10, where higher scores indicated greater knowledge. Attitudes toward evidence-based practice were measured using the Japanese version of the EBPAS [33]. The EBPAS comprises 15 items rated on a 0–4 Likert scale, and item scores were summed to yield a total score ranging from 0 to 60; higher scores reflected more favorable attitudes toward evidence-based practice. Internal consistency for the total EBPAS score in this study was acceptable (Cronbach’s α = 0.77). No changes to the pre-specified outcomes were made after trial commencement.

#### Data analysis

Baseline comparability between IG and CG was assessed using the chi-square test or Fisher’s exact test for categorical variables and independent t-tests for continuous variables, as appropriate. Because the primary and secondary outcome distributions were non-normal, between-group comparisons of CPD-Reaction Questionnaire scores, knowledge scores, and EBPAS scores at baseline, immediately post-intervention, and two weeks post-intervention were conducted using the Mann–Whitney U test. Missing outcome data were imputed using single imputation with the median. Analyses followed an intention-to-treat approach using the full analysis set. All tests were two-tailed with a significance threshold of p < .05. Effect sizes were reported as r = Z / √N, where Z is the standardized test statistic from the Mann– Whitney U test and N the total number of observations. Statistical analyses were performed with IBM SPSS Statistics, Version 29.0 [34].

#### Ethical considerations

This study was approved by the Medical Research Ethics Committee of Yokohama City University (2024-038), and informed consent was obtained from all subjects in writing. This RCT was approved by the appropriate ethical review committee and registered at the University Hospital Medical Information Network (UMIN) Center (No. UMIN000058251).

Although the trial was retrospectively registered in UMIN after participant enrollment had begun, the study protocol and outcome measures were established a priori and remained unchanged throughout the trial. The retrospective registration was due to administrative delay, and no selective outcome reporting occurred.

All participants were provided with written information regarding the purpose and methods of this study, protection of anonymity, voluntary participation, and how the results of this study would only be used for research purposes. The participants were informed that they were free to drop out, and they signed a consent form.

## Results

Figure 2 shows a flow diagram of study participant recruitment. Of the 158 participants, all were randomized: 79 in the IG and 79 in the CG. After allocation, five participants in the IG and seven in the CG dropped out. Two participants in the IG dropped out before the two week measurement, for a total of seven participants each from the IG and CG. Therefore, 72 participants in the IG and 72 participants in CG were included in the analysis. Seven participants in each group dropped out (5+2 in IG, 7 in CG). Reasons included schedule conflicts (n = 7) and illness or personal reasons (n = 7). There were no significant differences in baseline characteristics between completers and dropouts. No harm or unintended effects were observed during the study.

**Figure 2.**
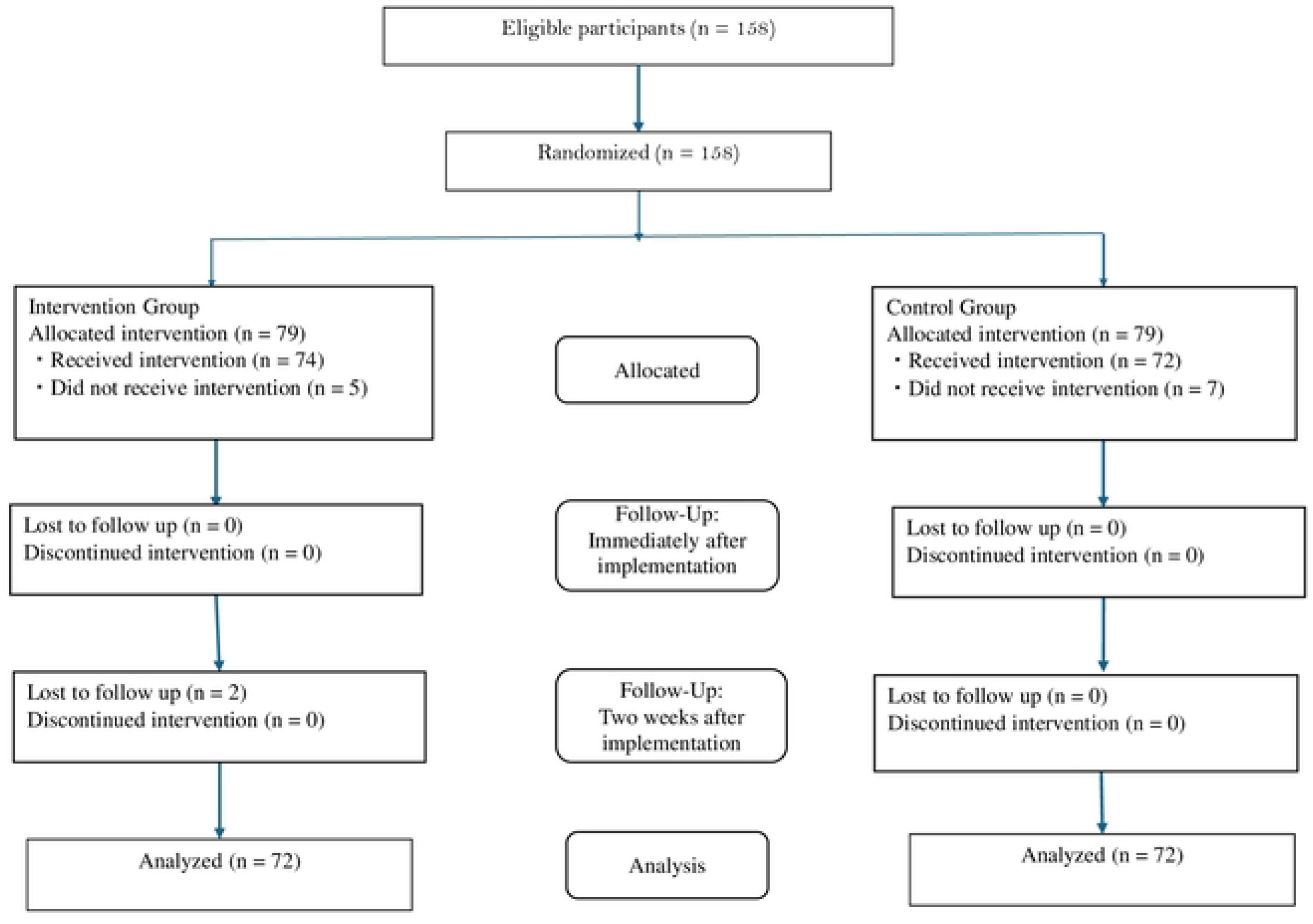
CONSORT flow diagram.

### Baseline participant characteristics

The attributes of participants in the intervention and control groups were compared (Table 2). No participant attributes differed significantly between the intervention and control groups (*p* > 0.05 for all items).

**Table. 2.** Demographics of participant.

| Demographics of participant | Intervention group(n=74) | Control group(n=72) | p-value |
| --- | --- | --- | --- |
| **Age** |  |  |  |
| 20s | 27 | 25 | .51 |
| 30s | 35 | 30 |  |
| 40s | 9 | 15 |  |
| 50s | 1 | 2 |  |
| 60s and above | 1 | 0 |  |
| **Year of experience as a midwife** |  |  |  |
| meas(SD) | 7.7(5.5) | 8.8(6.1) | .41 |
| **Years of experience in pregnancy health guidance** |  |  |  |
| mean(SD) | 5.4(4.2) | 6.9(5.9) | .77 |
| **Advanced midwife qualification** |  |  |  |
| yes | 6 | 11 | .14 |
| no | 68 | 61 |  |
| **Type of work** |  |  |  |
| University medical center | 15 | 6 | .15 |
| Hospital | 26 | 46 |  |
| Clinic | 14 | 16 |  |
| Midwifery clinic | 2 | 0 |  |
| Educational Institutions | 1 | 1 |  |
| Others | 6 | 3 |  |
| **Prior learning** |  |  |  |
| Yes | 2 | 1 | .61 |
| No | 72 | 71 |  |

Abbreviations: SD, standard deviation.
Prior learning: Whether there is midwifery care learning to prevent premature birth.
Chi-square test.: Age, Type of work.
Independent samples t-test.: Years of experience as a midwife, Years of experience in pregnancy health guidance.
Fisher's Chi-square test. : Advanced midwife qualification, Existing learning status.

### Effects of the education program on primary outcomes

The primary outcomes are shown in Table 3. There were no significant differences between the IG and CG in the intention to practice midwifery care to prevent premature births, as measured by the CPD-Reaction Questionnaire, before the implementation of the educational program (IG: 4.0 [IQR: 4.0–6.0], CG: 4.0 [IQR: 4.5–6.0]; p = .906, effect size = 0.01, small). Immediately after the program, the IG had significantly higher intention scores (6.5 [IQR: 5.5–7.0]) compared with the CG (5.5 [IQR: 5.0–6.5]; p = .005, effect size = 0.23, small). This difference persisted at the two-week follow-up (IG: 6.0 [IQR: 5.5–7.0], CG: 5.5 [IQR: 5.0–6.5]; p = .001, effect size = 0.27, medium), indicating that the program effectively increased participants’ intentions to engage in premature birth prevention practices

**Table. 3.** Comparison of IG and CG in CPD-Reaction Questionnare.

| Intention | Intervention group(74,<sup>a</sup>72) | Control group(72) | <sup>b</sup>HL diff (95% CI) | *p*-value | Effect size | <sup>c</sup> *r* |
| --- | --- | --- | --- | --- | --- | --- |
|  | MED [IQR] | MED [IQR] |  |  |  |  |
| Before the education program | 4.0 [4.0-6.0] | 4.0 [4.5-6.0] | 0.0 (-0.5 to 0.5) | .906 | 0.01 | small |
| Immediately after completion of the program | 6.5 [5.5-7.0] | 5.5 [5.0-6.5] | 0.5 ( 0.0 to 1.0) | .005 | 0.23 | small |
| 2 weeks after completion of the program | 6.0 [5.5-7.0] | 5.5 [5.0-6.5] | 0.5 ( 0.0 to 1.0) | .001 | 0.27 | small |
| **Social influence** |  |  |  |  |  |  |
|  | MED [IQR] | MED [IQR] |  |  |  |  |
| Before the education program | 4.6 [3.8-5.5] | 4.7 [3.6-5.4] | 0.0 (-0.3 to 0.5) | .793 | 0.02 | small |
| Immediately after completion of the program | 5.2 [4.3-5.9] | 5.1 [4.1-5.9] | 0.0 (-0.3 to 0.5) | .852 | 0.02 | small |
| 2 weeks after completion of the program | 6.6 [5.9-7.8] | 5.1 [4.4-5.9] | -1.7 (-2.1 to -1.3) | <.001 | 0.56 | large |
| **Beliefs about capabilities** |  |  |  |  |  |  |
|  | MED [IQR] | MED [IQR] |  |  |  |  |
| Before the education program | 3.8 [2.7-4.7] | 3.7 [2.4-4.3] | 0.3 (-0.3 to 0.7) | .260 | 0.09 | small |
| Immediately after completion of the program | 5.0 [4.3-6.0] | 4.3 [3.3-5.0] | 0.7 ( 0.3 to 1.0) | .002 | 0.26 | small |
| 2 weeks after completion of the program | 5.0 [4.0-5.7] | 4.3 [3.3-4.9] | 0.7 ( 0.3 to 1.0) | <.001 | 0.29 | small |
| **Moral norm** |  |  |  |  |  |  |
|  | MED [IQR] | MED [IQR] |  |  |  |  |
| Before the education program | 6.0 [4.9-7.0] | 6.0 [5.0-7.0] | 0.0 (-0.5 to 0.0) | .691 | 0.03 | small |
| Immediately after completion of the program | 6.5 [5.5-7.0] | 5.5 [4.5-6.0] | 1.0 (0.5 to 1.5) | <.001 | 0.45 | medium |
| 2 weeks after completion of the program | 6.5 [5.5-7.0] | 6.0 [5.5-7.0] | 0.0 (0.0 to 0.5) | .476 | 0.06 | small |
| **Beliefs about consequences** |  |  |  |  |  |  |
|  | MED [IQR] | MED [IQR] |  |  |  |  |
| Before the education program | 6.5 [6.0-7.0] | 7.0 [6.0-7.0] | 0.0 (-0.5 to 0.0) | .282 | 0.09 | small |
| Immediately after completion of the program | 7.0 [6.5-7.0] | 7.0 [6.0-7.0] | 0.0 (0.0 to 0.0) | .380 | 0.07 | small |
| 2 weeks after completion of the program | 7.0 [6.0-7.0] | 7.0 [6.0-7.0] | 0.0 (0.0 to 0.0) | .971 | 0.00 | small |

Abbreviations: MED, median IQR, interquartile range
note:The Mann–Whitney U test.: Intention, Social influence, Belief about capabilities, Moral norm, Beliefs about consequences
<sup>a</sup>2weeks after completion of the program <sup>b</sup>Hodges–Lehmann estimate of the median difference; CI = confidence interval.<sup>c</sup>*r* large >0.5, medium 0.3-0.5, small <0.3

Regarding social influence, no significant differences were found between the IG (4.6 [IQR: 3.8–5.5]) and the CG (4.7 [IQR: 3.6–5.4]) before the intervention (p = .793, effect size = 0.02, small), or immediately afterward (IG: 5.2 [IQR: 4.3–5.9], CG: 5.1 [IQR: 4.1–5.9]; p = .852, effect size = 0.02, small). However, two weeks after the intervention, the IG showed significantly higher social influence scores (6.6 [IQR: 5.9–7.8]) compared with the CG (5.1 [IQR: 4.4–5.9]; p < .001, effect size = 0.56, large), suggesting a delayed positive effect of the program on perceived social influence.

For beliefs about capabilities, there were no significant differences at baseline (IG: 3.8 [IQR: 2.7–4.7], CG: 3.7 [IQR: 2.4–4.3]; p = .260, effect size = 0.09, small). However, the IG had significantly higher scores than the CG both immediately after the intervention (IG: 5.0 [IQR: 4.3–6.0], CG: 4.3 [IQR: 3.3–5.0]; p=0.002, effect size = 0.26, small) and two weeks later (IG: 5.0 [IQR: 4.0–5.7], CG: 4.3 [IQR: 3.3–4.9]*; p* < .001, effect size = 0.29, small), indicating sustained improvements in participants’ perceived capabilities to provide premature birth prevention care.

Similarly, no differences were observed between groups in moral norms before the intervention (IG: 6.0 [IQR: 4.9–7.0], CG: 6.0 [IQR: 5.0–7.0]; p = .691, effect size = 0.03, small). The IG showed significantly higher moral norm scores immediately after the intervention (IG: 6.5 [IQR: 5.5–7.0], CG: 5.5 [IQR: 4.5–6.0]; p < .001, effect size = 0.45, medium), but this difference was no longer evident at the two-week follow-up (IG: 6.5 [IQR: 5.5–7.0], CG: 6.0 [IQR: 5.5–7.0]; p = .476, effect size = 0.06, small). These results suggest that the program temporarily enhanced the moral norms regarding premature birth prevention practices.

Finally, the beliefs about the consequences of midwifery care did not differ significantly between groups at any time point (before: IG: 6.5 [IQR: 6.0–7.0], CG: 7.0 [IQR: 6.0–7.0]; p = .282, effect size = 0.09, small; immediately after: IG: 7.0 [IQR: 6.5–7.0], CG: 7.0 [IQR: 6.0–7.0]; p = .380, effect size = 0.07, small; two weeks after: IG: 7.0 [IQR: 6.0– 7.0], CG: 7.0 [IQR: 6.0–7.0]; p = .971, effect size = 0.00, small), indicating that the program had no measurable effect on this construct.

### Effects of the education program on secondary outcomes

#### Knowledge of midwifery care to prevent premature birth

The secondary outcomes are presented in Table 4. There were no significant differences between the IG (5.0 [IQR: 3.0–6.0]) and the CG (5.0 [IQR: 3.0–6.0]) in knowledge of midwifery care to prevent premature birth before the implementation of the educational program (p = .721, effect size = 0.04, small). Immediately after implementation, the IG demonstrated significantly higher knowledge scores (9.0 [IQR: 9.0–10.0]) compared with the CG (6.0 [IQR: 6.0–7.0]; p < .001, effect size = 0.81, large). This significant difference was maintained two weeks later, with the IG scoring 9.0 (IQR: 8.0–10.0) and the CG scoring 6.0 (IQR: 4.3–7.0; p < .001, effect size = 0.72, large). These findings indicate that the educational program substantially improved participants’ knowledge of premature birth prevention, and this effect persisted for at least two weeks.

**Table. 4.** Comparison of IG and CG in knowledge scares and EBPAS.

| **knowledge scores** | Intervention group(74, <sup>a</sup>72) | Control group(72) | <sup>b</sup>HL diff (95% CI) | *p*-value | Effect size <sup>c</sup>*r* |  |
| --- | --- | --- | --- | --- | --- | --- |
|  | MED [IQR] | MED [IQR] |  |  |  |  |
| Before the education program | 5.0 [3.0-6.0] | 5.0 [3.0-6.0] | 0.0 (0.0 to 1.0) | .721 | 0.04 | small |
| Immediately after completion of the program | 9.0 [9.0-10.0] | 6.0 [6.0-7.0] | 3.0 (3.0 to 4.0) | <.001 | 0.81 | large |
| 2 weeks after completion of the program | 9.0 [8.0-10.0] | 6.0 [4.3-7.0] | 3.0 (2.0 to 3.0) | <.001 | 0.72 | large |
| **EBPAS** |  |  |  |  |  |  |
|  | MED [IQR] | MED [IQR] |  |  |  |  |
| Before the education program | 42.0 [39.0-47.0] | 45.0 [40.0-49.8] | - 2.0 (- 4.0 to 0.0) | .102 | 0.14 | small |
| Immediately after completion of the program | 47.0 [42.0-50.0] | 46.5 [41.0-51.0] | 0.0 (- 2.0 to 2.0) | .870 | 0.02 | small |
| 2 weeks after completion of the program | 44.0 [39.0-49.0] | 43.0 [39.0-47.0] | 1.0 (- 1.0 to 3.0) | .362 | 0.09 | small |

Abbreviations:MED, median IQR, interquartile range
note:The Mann–Whitney U test.: Knowledge score; EBPAS <sup>a</sup>2weeks after completion of the program
<sup>b</sup>Hodges–Lehmann estimate of the median difference; CI = confidence interval. <sup>c</sup>*r* large >0.5, medium 0.3-0.5, small <0.3

#### Evidence-based practice attitude scale

There were no significant differences in EBPAS scores between the groups at any time point. Before the implementation, the IG scored 42.0 (IQR: 39.0–47.0) and the CG scored 45.0 (IQR: 40.0–49.8; p = .142, effect size = 0.14, small). Immediately after the program, the IG scored 47.0 (IQR: 42.0–50.0) and the CG scored 46.5 (IQR: 41.0–51.0; p = .870, effect size = 0.02, small). Two weeks after implementation, the IG scored 44.0 (IQR: 39.0–49.0) and the CG scored 43.0 (IQR: 39.0–47.0; p = .362, effect size = 0.09, small). These results suggest that the educational program had no measurable effect on participants’ attitudes toward evidence-based practice.

## Discussion

### Effects of the education program

The results of this study showed that midwives who took the online interactive education program to prevent premature birth had significantly higher intention and knowledge to practice midwifery care to prevent premature births immediately after and two weeks after the program, compared with midwives who read a pamphlet containing evidence. Thus, our hypotheses were partially supported.

A systematic review of healthcare professional intentions and behavior found intention to be a strong influencer of behavior, reporting 25.6%–34.0% of the explained variance in TPB behavior [23]. Another study on an educational program for medical professionals using the TPB showed that intentions improved following intervention, and as a result, intentions were put into behavior [35]. This study investigated the intentions to implement midwifery care to prevent premature birth, not the actual practice. However, it is expected that intentions will translate into practice.

A meaningful result of this study was that intentions to practice midwifery care to prevent premature birth and beliefs about capabilities related to intentions increased immediately after the educational program intervention and were maintained two weeks later. Some authors have reported stronger effects of beliefs about capabilities on intentions [36, 37], and it is possible that the increases in beliefs about capabilities also influenced intention in this program. In the CPD-Reaction Questionnaire, beliefs about capabilities (a construct conceptually related to perceived behavioral control or self-efficacy in the TPB) represent the belief that one can perform an action and reflect a sense of self-confidence and self-control.

In the educational program we developed, midwives engaged in dialogue with each other to discuss and encourage each other regarding the difficulties and solutions they faced when practicing evidence-based midwifery care to prevent premature births. In addition, the facilitator provided real-time feedback on midwives’ opinions. According to Bandura [38], a person’s self-efficacy can be influenced by observing others’ success in their actions and through encouragement. Therefore, the implementation of an interactive program that provides an opportunity to discuss practical midwifery care to prevent premature birth may have been effective in encouraging midwives to consider the midwifery care they can provide in their settings to prevent premature births and to increase their belief in their capabilities.

In addition, the results of the educational program showed that the knowledge of midwifery care to prevent premature birth increased significantly in the IG compared with the CG and was maintained even after two weeks. Behavioral change requires the possession of knowledge and skills [39], and knowledge acquisition provides the foundation for implementing evidence-based midwifery care to prevent premature birth. We believe that barriers to EBP implementation among midwives include the lack of knowledge and skills, meaning that increased knowledge will promote evidence-based midwifery care. However, there was no difference in evidence-based attitudes between the IG and CG before and after the intervention or at the two-week mark. The educational program we developed set pre- and post-tasks and asked midwives to review their antenatal care with the hope of changing their attitudes by making them aware of the shortcomings in midwifery care practice to prevent premature birth. Midwives are experts in supporting pregnancy, childbirth, and childcare, and the philosophy of midwifery care is consistent with EBP [40]. Previous studies have shown that midwives have a positive attitude toward EBP [41] and, regardless of the presence of intervention, both the IG and CG positively assessed EBP, which may explain the lack of differences.

Notably, the absence of differences in evidence-based attitudes between the intervention and control groups may also be attributed to a potential ceiling effect, as the participating midwives may have already had high baseline scores on the EBPAS, leaving little room for further improvement after the intervention [42, 43].

### Significance of the educational program for preventing premature birth

To the best of our knowledge, this may be one of the first midwifery education programs worldwide to specifically focus on preventing premature births. In addition to providing knowledge-based lectures, the program was designed to enhance midwives’ intentions to practice preventive midwifery care and to promote behavioral change based on the TPB. Although the premature birth rate in Japan is lower than in many other countries, the long-term administration of β-agonists has traditionally been used to prevent premature birth, such as ritodrine hydrochloride [44]. Approximately 30% of pregnant women diagnosed with threatened premature birth in Japan were prescribed ritodrine hydrochloride for 28 days or longer [45]. While β-agonists suppress uterine contractions, their prolonged use carries a high risk of serious adverse events for both the mother and fetus [46]. Therefore, to shift from the conventional pharmacological approach toward non-pharmacological, preventive strategies are urgently needed.

The strength of this educational program is that it identified effective, evidence-based interventions to prevent premature birth and educated midwives on comprehensive strategies to improve the physical, psychological, and environmental well-being of pregnant women. Previous training programs for midwives have tended to focus on single topics, such as nutrition during pregnancy [47] or screening for intimate partner violence [48]. However, the risk factors for premature birth are diverse and require multifaceted interventions. This new program integrates multiple evidence-based approaches into a single framework, offering midwives practical strategies for addressing the body, mind, and environment of pregnant women. In developed countries such as Japan, excessive medical interventions are often criticized for contributing to poor maternal outcomes [19]. Midwives need to recognize that maintaining a healthy pregnancy through evidence-based preventive care, rather than relying solely on drug therapy, can help reduce premature births. By equipping midwives with practical knowledge and fostering behavioral change, this program represents an innovative approach to transforming midwifery care from treatment to prevention.

### Limitations and directions for future research

This study has several limitations that should be considered when interpreting the findings. First, it applied the TPB to increase midwives’ intention to provide midwifery care to prevent preterm birth. Although this intention could potentially translate into actual practice, the extent to which participants implemented the intended behaviors was not measured. Future research should examine the relationship between intention and actual practice, as well as the link between midwifery practice and reductions in the preterm birth rate. Second, as the effects were assessed only two weeks after the educational program, its long-term effects remain unknown. Future studies should evaluate the sustainability of midwives’ practice to prevent preterm birth after the program. Third, the participants and researchers were not blinded, which may have introduced performance and detection bias and affected internal validity.

## Conclusions

This randomized controlled trial demonstrated that an online interactive midwifery education program based on the TPB effectively improved midwives’ intention and knowledge regarding premature birth prevention. By fostering intention—an established precursor to behavior change—and equipping midwives with practical, evidence-based strategies, the program has the potential to shift midwifery practice from treatment-oriented to prevention-focused care. This approach may contribute to reducing the risk of premature birth and improving maternal and neonatal health outcomes. Further large-scale studies with diverse populations and longer follow-up periods are warranted to evaluate the long-term effectiveness and generalizability of this educational program.

## Data availability statement

All relevant data are provided within the paper and the supporting information file.

## Data Availability

All relevant data are within the manuscript and its Supporting Information files.

## Acknowledgments

The authors sincerely thank the nursing professionals at the collaborating facilities for assisting with this study. This study was supported by the Yamaji Fumiko Specialized Nursing Education Research Grant Fund.

## Author contributions

Masaki Kidera: Conceptualization, Methodology, Investigation, Formal analysis, Data curation, Project administration, Funding acquisition, Writing – original draft, Writing – review & editing.

Shoko Takeuchi: Methodology, Data curation, Formal analysis, Writing – review & editing. Eriko Shinohara: Methodology, Data curation, Formal analysis, Writing – review & editing. Kazumi Kubota: Methodology, Formal analysis.

Sachiyo Nakamura: Conceptualization, Methodology, Supervision, Formal analysis, Writing – review & editing.All authors read and approved the final manuscript.

